# Patient-Specific EEG Baseline Establishment Using the E-norms Method for Pediatric Seizure Detection Without Labeled Training Data

**DOI:** 10.64898/2026.07.13.26357876

**Authors:** Joe F Jabre

## Abstract

The aim of this work is to validate patient-specific EEG baseline establishment using the e-norms method as a screening and retrospective-review tool for seizure detection in pediatric epilepsy. The method was applied to 247 seizure-free EEG recordings (263.92 hours) from 10 patients in the CHB-MIT Scalp EEG Database (ages 3–18). A composite stability metric combining first-derivative dynamics, spectral entropy, variance, and line length was computed per 2-second epoch across 23 channels. Patient-specific detection thresholds were derived from each patient’s seizure-free baseline using a weighted statistical procedure. Performance was validated against 72 expert-annotated seizures (2,705 epochs) across 62 seizure files, with durations spanning 6 to 264 seconds (44-fold range). The results show that detection achieved 94.4% event-level sensitivity (68 of 72 seizures; 95% CI 86.6–97.8%) and 81.5% epoch-level sensitivity (2,204 of 2,705 epochs; 95% CI 80.0– 82.9%). Eight of ten patients achieved 100% event-level sensitivity with epoch-level sensitivity ranging from 58.7% to 100.0%. Two patients showed partial event-level failures (CHB-15: 17 of 20; CHB-18: 5 of 6), with the four missed events attributable to two characterizable failure modes (Section 4.5). Patient-specific thresholds ranged from 4.06 to 4.81 (mean 4.51 ± 0.25); threshold variation did not correlate reliably with age or sex, confirming that no universal threshold could achieve comparable performance. Detection margins ranged from 0.88 to 1.24 times. Patient-specific e-norms achieves 94.4% event-level sensitivity for pediatric EEG seizure detection without requiring labeled seizure training data, exceeding published human expert inter-rater agreement (50–76%) and recent automated approaches in adult cohorts using behind-the-ear EEG and wearable ECG. Two characterizable failure modes account for the four missed events and inform appropriate clinical use. As a high-sensitivity screening tool complementary to real-time alarm systems, the method is ready for adult validation, prospective deployment, and head-to-head benchmarking.

## 1. Introduction

The interpretation of diagnostic studies in clinical neurophysiology depends fundamentally on appropriate normal values. Most EEG laboratories use age-appropriate, population-derived normative databases — values typically obtained from external cohorts using different equipment and electrode configurations, sometimes collected decades earlier.

Based on current guidelines and literature, including the American Clinical Neurophysiology Society (ACNS) [1], [2], [3], routine clinical EEG interpretation is grounded in standardized, age- and state-adjusted reference patterns, whereas patient-specific normative approaches are primarily reported in specialized QEEG or research settings.

In this work, I describe the use of the e-norms method I developed and published in 2015 [4], originally to derive EMG and nerve conduction studies normal values. The method identifies normal values from mixed datasets containing both normal and abnormal studies through their characteristic clustering behavior [5] — an ‘a posteriori’ approach that extracts normative values from existing patient data, eliminating the need to recruit healthy reference cohorts.

Since its first deployment for EMG and nerve conduction studies, the e-norms method has been validated against other methods using the same a posteriori construct [6] and applied across multiple domains including body mass index [7], nerve and muscle ultrasounds [8], ophthalmology [9], orthopedics [10], acute phase inflammatory markers [11], and pediatric neurophysiology [12], [13].

Despite this breadth of application, e-norms has not been applied to neurophysiological time-series data such as EEG. The challenge is fundamental: peripheral electrophysiology and ultrasound yield discrete measurements amenable to direct sorting and plateau identification, whereas raw EEG signals comprise multidimensional waveforms that cannot be sorted in their native form.

The aim of this work is to describe the first application of the e-norms methodology to neurophysiological time-series data, specifically to patient-specific EEG baseline characterization for seizure detection.

Epilepsy affects approximately 50 million people worldwide [14], and accurate seizure detection in continuous EEG is essential to clinical decision-making in epilepsy monitoring units (EMUs) and intensive care settings. Expert visual review remains labor-intensive and inconsistent, with reported sensitivity often in the 20– 50% range [15], [16]. In pediatric populations, the detection challenge is particularly acute [17].

Current automated approaches rely on either population-based normative data or supervised machine learning trained on labeled seizures [18], [19]. Both face limitations in pediatric epilepsy: inter-individual variation in normal EEG patterns [20], scarce labeled training data, and poor generalizability across patient subsets [21].

Deriving detection thresholds from each patient’s own seizure-free recordings would, in principle, capture individual variation while eliminating the need for labeled seizure training data — particularly attractive in pediatric epilepsy, where baseline EEG patterns vary substantially with age and developmental stage [22],[23]. The barrier has been practical: systematic baseline characterization requires clinical, statistical, and computational expertise that single investigators rarely possess.

This work extends the e-norms methodology to multi-channel EEG time-series data through a composite stability metric, validated on the CHB-MIT database [24]. The methodology is positioned as a high-sensitivity screening and retrospective-review tool, complementary to real-time alarm systems. Patient-specific baselines from seizure-free recordings were derived via duration × cohesion-weighted thresholds, and seizure detection was validated across the full pediatric developmental age range (Table 1).

**Table 1.** Patient Cohort Demographics and Baseline Characteristics.

| Patient | Age (yr) | Sex | Baseline Files | Baseline Hours | Threshold |
| --- | --- | --- | --- | --- | --- |
| CHB-10 | 3-7 | M | 18 | 36.01 | 4.8036 |
| CHB-08 | 3-7 | M | 15 | 15.01 | 4.8139 |
| CHB-16 | 3-7 | F | 13 | 13.00 | 4.4258 |
| CHB-01 | 8-12 | F | 34 | 32.91 | 4.6454 |
| CHB-02 | 8-12 | M | 33 | 33.00 | 4.6110 |
| CHB-17 | 8-12 | F | 18 | 18.00 | 4.6439 |
| CHB-21 | 13-17 | F | 29 | 29.00 | 4.3977 |
| CHB-03 | 13-17 | F | 31 | 31.00 | 4.5729 |
| CHB-15 | 13-17 | M | 26 | 26.00 | 4.0589 |
| CHB-18 | 18-22 | F | 30 | 30.00 | 4.1676 |
| **Combined** | **3–18** | **6F/4M** | **247** | **263.92** | **4.514 ± 0.251** |

*Threshold computed per Section 2.3.*
*Combined threshold reported as mean ± SD across the 10 patient-specific values*

## 2. Materials and Methods

### 2.1 Database and patient selection

The CHB-MIT Scalp EEG Database, publicly available through PhysioNet (https://physionet.org/content/chbmit/1.0.0/), contains continuous scalp EEG recordings from 22 pediatric patients with intractable seizures monitored at Children’s Hospital Boston (chb). This database is one of the most widely used benchmarks for seizure detection algorithm development [25]. Recordings use the international 10–20 electrode system in a 23-channel bipolar montage sampled at 256 Hz, stored as European Data Format (EDF) files.

Ten patients were selected for validation, spanning the full pediatric-to-young adult developmental range (ages 3–18 years; 6 female, 4 male). Patient selection prioritized age diversity, balanced sex representation, and varied recording and seizure characteristics. Across all 10 patients, the dataset comprised 309 EDF files (247 seizure-free baseline files and 62 seizure-containing files; some files contain multiple seizures), representing 263.92 hours of baseline EEG and 72 expert-annotated seizure events containing 2,705 seizure epochs. Per-patient demographics, baseline characteristics, and seizure counts are summarized in Table 1.

### 2.2 Composite stability metric

Traditional e-norms methodology applies directly to single-value measurements (e.g., nerve conduction amplitudes). Raw EEG signals present fundamental challenges: multidimensional data structure (23 channels × continuous waveforms) and context-dependent voltage values that cannot be meaningfully sorted. To address this, we developed a two-level hierarchical architecture using an engineered composite stability metric (Figure 1).

**Fig. 1:**
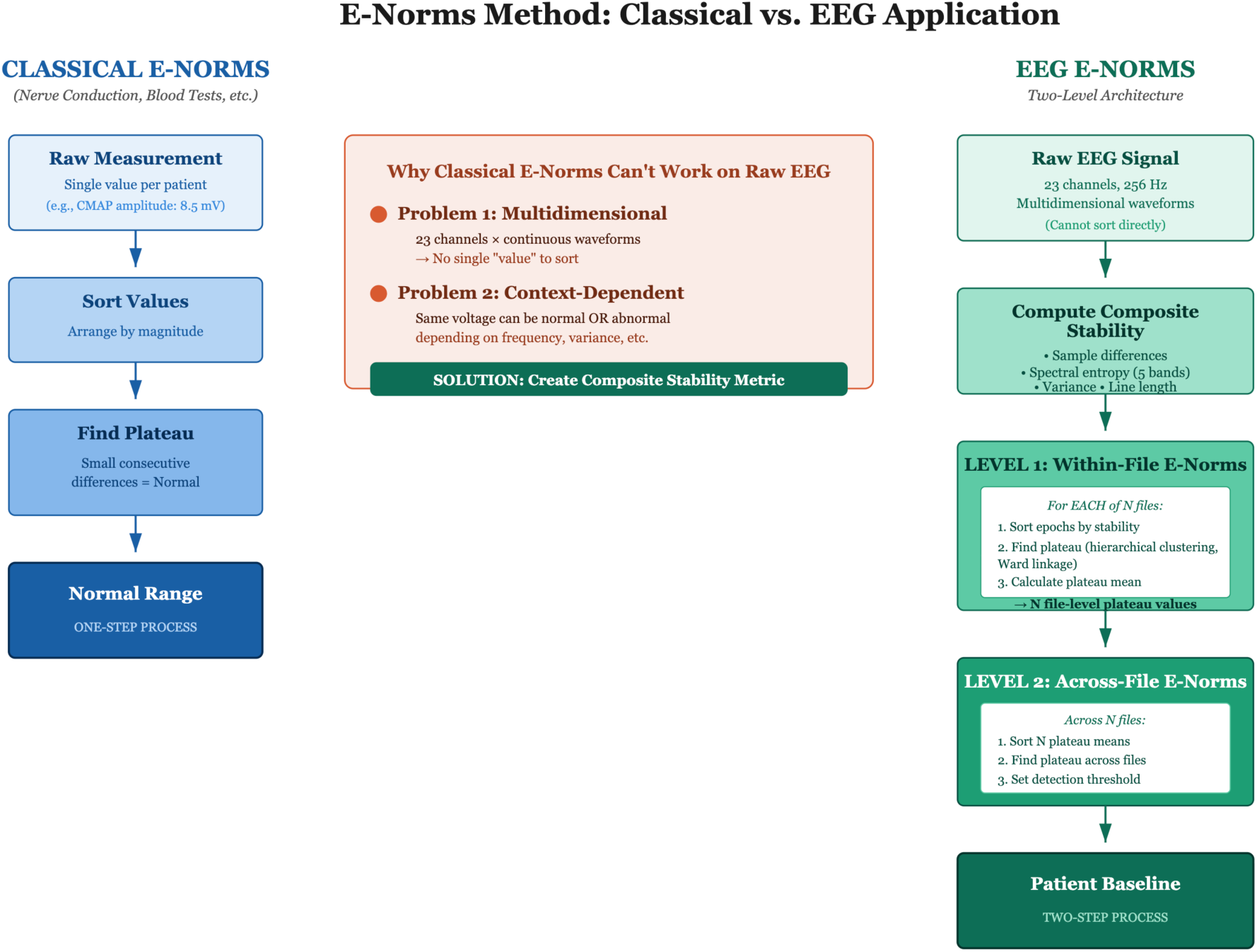
Methodological extension of e-norms from single measurements to composite features.

Each EEG recording was segmented into non-overlapping 2-second epochs. Composite stability was computed per epoch across all 23 channels as a dimensionless metric aggregating four signal characteristics with fixed weights:

Composite_stability = 0.3 · (diff_mean / diff_std) + 0.3 · spectral_entropy + 0.2 · log1p(variance) + 0.2 · log1p(line_length)

The four components capture distinct aspects of EEG signal stability: diff_mean / diff_std reflects moment-to-moment voltage dynamics; spectral_entropy is the mean Shannon entropy across the standard EEG frequency bands (delta 0.5–4 Hz, theta 4– 8 Hz, alpha 8–13 Hz, beta 13–30 Hz, gamma 30–50 Hz); variance reflects signal power; and line length captures the total path length of the signal trajectory. The log1p transformation on variance and line length brings them onto a comparable scale with the other terms (Figure 2).

**Fig. 2:**
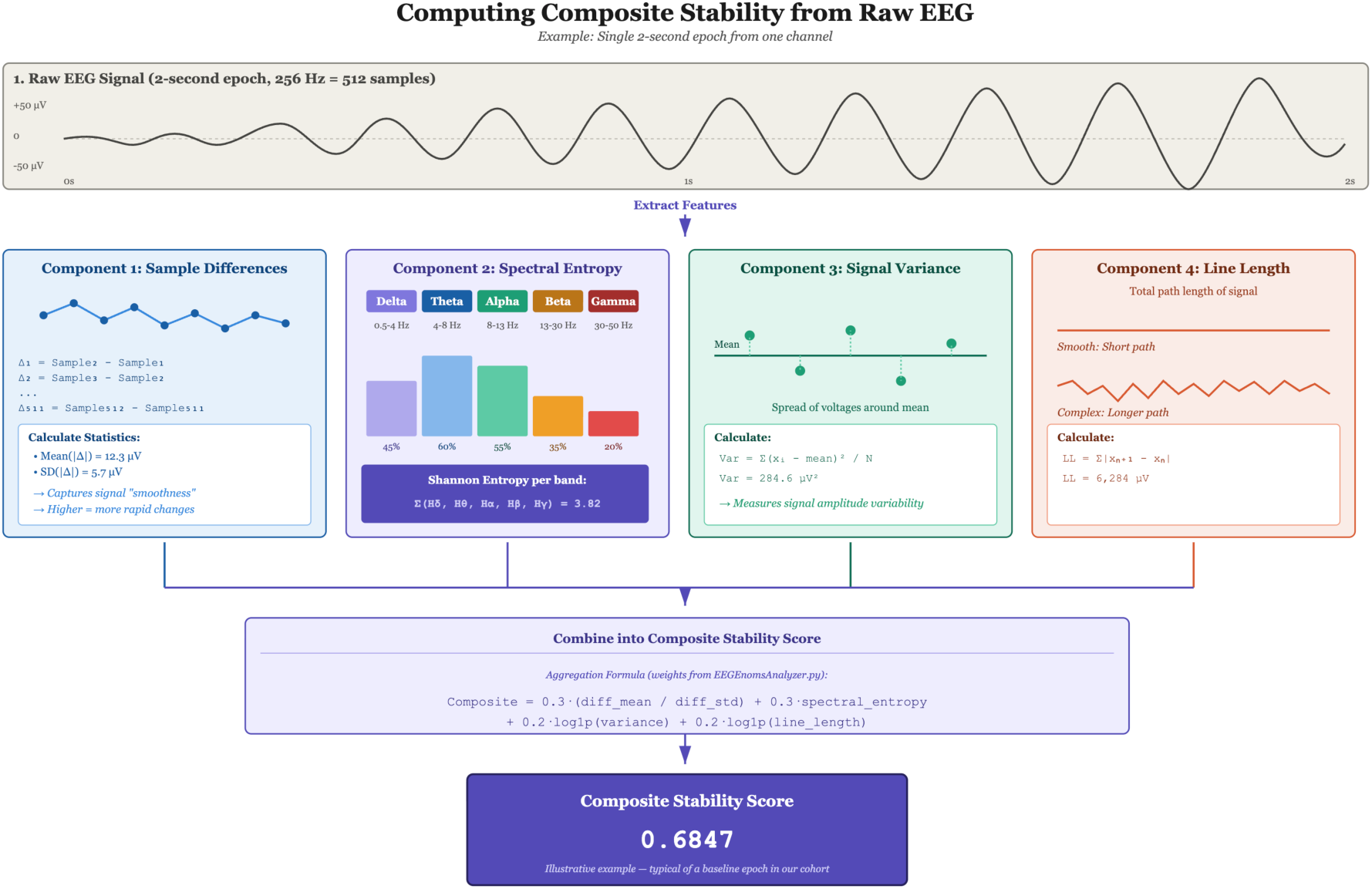
Derivation of composite stability metric from raw EEG signals.

Within each file, epochs were classified as normal (belonging to the dominant stability plateau) or abnormal (outliers and artifacts) using hierarchical agglomerative clustering with Ward linkage. The number of clusters (between 3 and 5) was selected automatically by maximizing silhouette score, and the plateau cluster was identified as the largest cluster with the lowest mean composite stability. For each file, the following were calculated: (1) plateau mean and standard deviation among normal epochs, (2) cohesion (percentage of epochs classified as normal), and (3) coefficient of variation (CV = SD/mean × 100%).

### 2.3 Patient-specific baseline establishment and threshold derivation

For each patient, all seizure-free EDF files were processed through the composite stability pipeline to establish a patient-specific normal baseline. Across the 10 patients, the number of baseline files per patient ranged from 13 (CHB-16) to 34 (CHB-01), with a median of 27.5 files and 29.5 hours of baseline EEG per patient (cohort total: 247 files, 263.92 hours). Each file yielded a plateau mean (m_i), plateau standard deviation (s_i), cohesion (c_i, the percentage of epochs classified as normal), and recording duration in minutes (d_i), characterizing its contribution to the patient’s normal EEG range.

File-level plateau values were consolidated into a single patient-specific threshold using duration × cohesion weighting. For each file i, the weight was defined as w_i = d_i × (c_i / 100), prioritizing long, stable recordings while down-weighting short or artifact-contaminated segments. The weighted mean and weighted standard deviation across the patient’s baseline files were computed as:

Weighted_mean = Σ (w_i / Σw) · m_i

Weighted_SD = √[ Σ (w_i / Σw) · (s_i² + (m_i − Weighted_mean)²) ]

The combined weighted SD formula incorporates both within-file variance (s_i²) and between-file variance ((m_i − Weighted_mean)²), giving a single estimate of plateau dispersion that reflects the patient’s overall baseline characteristics rather than any single recording.

The patient-specific detection threshold was then calculated as:

Threshold = Weighted_mean + 1.0 × Weighted_SD

The multiplier was held constant across all 10 patients; only Weighted_mean and Weighted_SD vary by patient, applying a uniform methodology to individualized baselines.

### 2.4 AI-Enabled collaborative analysis paradigm

Analyses were conducted through human-AI collaboration using Claude (Anthropic). The investigator specified clinical hypotheses, methodological framework, cohort composition, and result interpretation; the AI implemented computational analyses, performed statistical calculations, and drafted code that was reviewed and accepted by the investigator before application to the data. All final interpretations, clinical decisions, and conclusions remained the responsibility of the investigator.

### 2.5 Seizure detection validation

Seizure detection was validated on 62 EDF files containing 72 expert-annotated seizure events across all 10 patients. Seizure onset and offset times were obtained from the CHB-MIT database annotations, originally provided by expert epileptologists at Children’s Hospital Boston. For each seizure file, composite stability was computed for every 2-second epoch (Section 2.2) and compared to the patient-specific threshold (Section 2.3). Epochs with composite stability exceeding threshold were classified as seizure-positive.

Two complementary sensitivity metrics were defined. Event-level sensitivity was the proportion of seizure events with at least one detected epoch within the annotated interval — addressing whether each seizure was identified at all, the primary requirement for a screening or retrospective-review tool. Epoch-level sensitivity was the proportion of all 2-second epochs within annotated seizure intervals that exceeded the patient-specific threshold — additionally indicating whether the entire seizure period was flagged, relevant to downstream applications such as seizure burden quantification.

The validation set spanned a wide range of clinical scenarios. Seizure files ranged from single-seizure recordings to files containing up to 5 seizures (CHB-15 file 54). Seizure durations ranged from 6 seconds (4 epochs, CHB-16) to 264 seconds (133 epochs, CHB-08), a 44-fold range spanning brief electrographic seizures to prolonged ictal events.

### 2.6 Statistical analysis

Both event-level and epoch-level sensitivities (definitions in Section 2.5) are reported with 95% binomial confidence intervals using the Wilson score method. Cohort-level sensitivities were computed by summing detected and total counts across all patients before taking the ratio (i.e., epoch-weighted, not patient-weighted). Patient-specific thresholds were summarized with descriptive statistics (mean, SD, range, minimum, maximum). Detection margins were quantified per seizure as the ratio of maximum composite stability during the seizure interval to the patient-specific threshold. All analyses were performed using Python 3.9 with NumPy, Pandas, SciPy, and pyedflib.

## 3. Results

### 3.1 Patient cohort and baseline characteristics

Ten patients were analyzed, spanning ages 3 to 18 years with balanced sex distribution (6 female, 4 male). The cohort ranged from toddlers through adolescents approaching adulthood, distributed across the age bands shown in Table 1. A total of 247 seizure-free baseline files were processed, comprising 263.92 hours of EEG. Baseline sizes ranged from 13 files / 13.00 hours (CHB-16) to 34 files / 32.91 hours (CHB-01), with a median of 27.5 files and 29.5 hours per patient. Patient-specific thresholds ranged from 4.06 (CHB-15) to 4.81 (CHB-08), with mean 4.51 ± 0.25 (Table 1).

Although patients spanned a wide age range, threshold variability across the cohort showed no monotonic relationship with age or sex (Section 3.3), supporting the patient-specific approach over any age- or demographic-based normative scheme.

### 3.2 Seizure detection performance

Across the 10-patient cohort, 68 of 72 expert-annotated seizures were detected (event-level sensitivity 94.4%; 95% CI 86.6–97.8%) and 2,204 of 2,705 seizure epochs (epoch-level sensitivity 81.5%; 95% CI 80.0–82.9%) (Table 2). Eight of ten patients reached 100% event-level sensitivity, with epoch-level sensitivity ranging from 58.7% (CHB-17) to 100.0% (CHB-02). Two patients showed partial event-level failures: CHB-15 detected 17 of 20 seizures (85.0% event, 67.2% epoch), and CHB-18 detected 5 of 6 (83.3% event, 86.0% epoch). The four missed events are characterized by failure mode in Section 3.5.

**Table 2.**
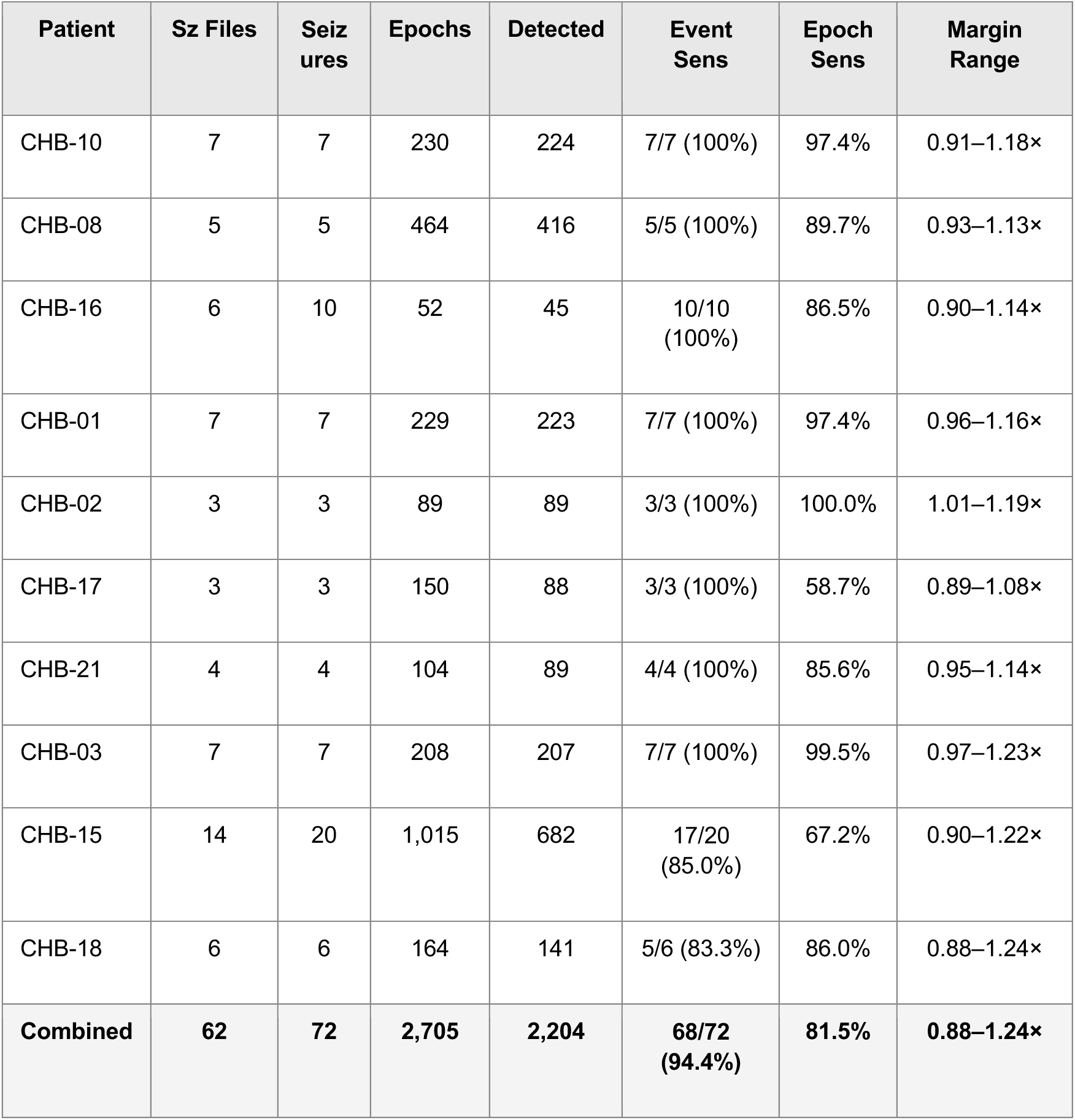
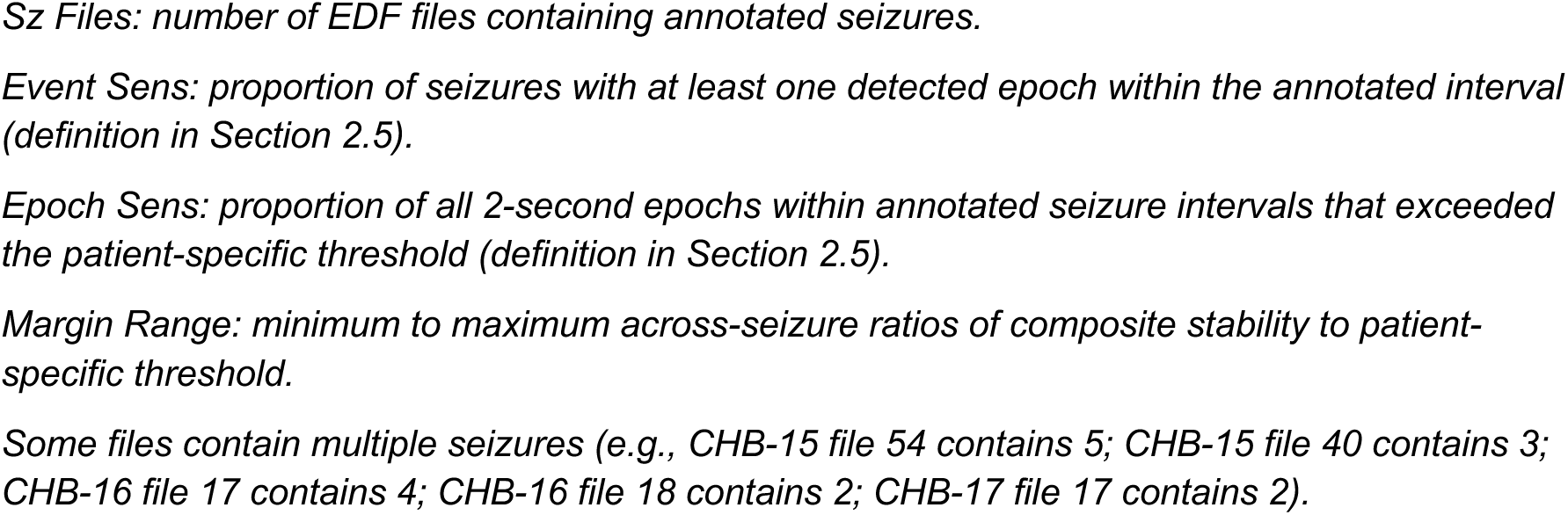
Seizure Detection Performance by Patient.

| Patient | Sz Files | Seizures | Epochs | Detected | Event Sens | Epoch Sens | Margin Range |
| --- | --- | --- | --- | --- | --- | --- | --- |
| CHB-10 | 7 | 7 | 230 | 224 | 7/7 (100%) | 97.4% | 0.91–1.18× |
| CHB-08 | 5 | 5 | 464 | 416 | 5/5 (100%) | 89.7% | 0.93–1.13× |
| CHB-16 | 6 | 10 | 52 | 45 | 10/10 (100%) | 86.5% | 0.90–1.14× |
| CHB-01 | 7 | 7 | 229 | 223 | 7/7 (100%) | 97.4% | 0.96–1.16× |
| CHB-02 | 3 | 3 | 89 | 89 | 3/3 (100%) | 100.0% | 1.01–1.19× |
| CHB-17 | 3 | 3 | 150 | 88 | 3/3 (100%) | 58.7% | 0.89–1.08× |
| CHB-21 | 4 | 4 | 104 | 89 | 4/4 (100%) | 85.6% | 0.95–1.14× |
| CHB-03 | 7 | 7 | 208 | 207 | 7/7 (100%) | 99.5% | 0.97–1.23× |
| CHB-15 | 14 | 20 | 1,015 | 682 | 17/20 (85.0%) | 67.2% | 0.90–1.22× |
| CHB-18 | 6 | 6 | 164 | 141 | 5/6 (83.3%) | 86.0% | 0.88–1.24× |
| **Combined** | **62** | **72** | **2,705** | **2,204** | **68/72 (94.4%)** | **81.5%** | **0.88–1.24×** |

*Sz Files: number of EDF files containing annotated seizures.*
*Event Sens: proportion of seizures with at least one detected epoch within the annotated interval (definition in Section 2.5).*
*Epoch Sens: proportion of all 2-second epochs within annotated seizure intervals that exceeded the patient-specific threshold (definition in Section 2.5).*
*Margin Range: minimum to maximum across-seizure ratios of composite stability to patient-specific threshold.*
*Some files contain multiple seizures (e.g., CHB-15 file 54 contains 5; CHB-15 file 40 contains 3; CHB-16 file 17 contains 4; CHB-16 file 18 contains 2; CHB-17 file 17 contains 2).*

Detection margins were modest by design. Per-seizure ratios of maximum composite stability to the patient-specific threshold ranged from 0.88 to 1.24 across the cohort, with both extremes observed in CHB-18 (smallest minimum margin 0.884×, largest maximum margin 1.242×). Margins reflect the patient-specific calibration of composite stability against each patient’s own baseline plateau: each patient’s seizures cross threshold by an amount proportional to that patient’s plateau dispersion, rather than with universal headroom. The cohort encompassed seizures across the full clinical duration spectrum: the shortest was 6 seconds (CHB-16, files 16 and 17) and the longest was 264 seconds (CHB-08 file 21), a 44-fold range. Both 6-second seizures were detected at event level, demonstrating that the methodology is not constrained to long seizures.

### 3.3 Patient-specific threshold analysis

Patient-specific thresholds ranged from 4.06 (CHB-15) to 4.81 (CHB-08), a 0.75 spread with mean 4.51 ± 0.25. Critically, threshold variation did not track reliably with age or sex. The two youngest patients (CHB-08, CHB-10) and the two oldest (CHB-15, CHB-18) clustered at the threshold extremes, suggesting an apparent age trend; however, the cohort as a whole showed no monotonic ordering — CHB-16 (threshold 4.43) is lower than CHB-01 (4.65) despite being younger, and CHB-21 (4.40) is lower than CHB-03 (4.57) despite being in the same age band. Same-age opposite-sex pairs (CHB-01 and CHB-02, in the same age band) had nearly identical thresholds (4.65 versus 4.61), while same-sex same-age-bracket patients (CHB-21 vs CHB-03) differed by 0.17. These observations confirm that detection thresholds are patient-specific properties that cannot be reliably predicted from demographic variables, validating the patient-specific e-norms approach.

Across this inter-individual threshold variation, patient-specific calibration achieved 100% event-level sensitivity in 8 of 10 patients and 94.4% across the cohort (Tables 1 and 2). The two partial-failure patients (CHB-15, CHB-18) had thresholds at the lower end of the cohort range (4.06 and 4.17), reflecting tighter baseline plateau dispersion rather than methodology adjustment; the threshold-derivation procedure applied uniformly to all 10 patients.

### 3.4 Robustness to recording variation

Although all CHB-MIT recordings follow the same 23-channel bipolar montage at 256 Hz, the dataset contains structural variation that the automated pipeline handled transparently. CHB-17 was recorded across three distinct sessions (chb17a, chb17b, chb17c) with separate file numbering for each session; baseline derivation pooled across all three sessions yielded a single patient-specific threshold (4.6439) that achieved 100% event-level sensitivity (3 of 3 seizures detected) across the patient’s seizure files. Several patients had non-contiguous file numbering reflecting clinical recording schedules (e.g., CHB-10 with files numbered up to 89 and CHB-15 with files numbered up to 62), which the pipeline handled without modification. CHB-02 was reanalyzed end-to-end on a fresh download from PhysioNet under the finalized pipeline, demonstrating that the methodology produces consistent results when re- applied from raw EDF to final detection. Across all 10 patients, no manual intervention was required at any stage: file discovery, baseline derivation, threshold computation, and seizure detection were fully automated.

### 3.5 Characterization of missed events

The four missed seizures distributed unevenly: three in CHB-15 (chb15_06, chb15_10, chb15_17) and one in CHB-18 (chb18_30). Quantitative characterization identified two distinct failure modes, each accounting for two missed events.

The first failure mode, observed in chb15_06 and chb15_10, reflects state-dependent baseline variability. CHB-15’s per-file plateau values varied substantially across 26 baseline files (range 3.73–4.17, across-file SD 0.10 — comparable to within-file dispersion). For both missed events, local plateau values in surrounding files (within ±5 files chronologically) averaged 3.82, below the cohort-aggregated plateau mean of 3.89. A supplementary investigation derived alternative thresholds from these local windows using the §2.3 procedure. Under local thresholding, chb15_06 was detected at 6 of 63 seizure epochs (versus 0 at the cohort threshold) and chb15_10 at 4 of 16 (versus 0). Both events were recoverable at event level.

The second failure mode, observed in chb15_17 and chb18_30, reflects ictal patterns that do not exceed the patient’s baseline plateau range. In chb15_17, maximum composite stability during the seizure (3.96) was only 0.4 SD above plateau mean (3.89); local thresholding also failed to recover this event (0 of 19 epochs at local threshold 4.04). In chb18_30, stability was at or below plateau throughout (mean 3.86, minimum 3.68; plateau mean 3.90). Implications and possible mechanisms for events of this character are addressed in Section 4.5.

## 4. Discussion

This 10-patient validation demonstrates that patient-specific e-norms methodology achieves 94.4% event-level sensitivity for automated EEG seizure detection across the full pediatric developmental range, without requiring labeled seizure training data. The validation tests the methodology across diverse clinical scenarios — toddler through near-adult EEG and seizure durations spanning a 44-fold range. The key methodological innovation — applying e-norms to engineered composite features rather than directly measured variables, a feature that can be termed e-norms+ — successfully generalizes the approach from single-measurement applications to complex neurophysiological time-series data.

### 4.1 Patient-specific vs population-based approaches

A central finding is that detection thresholds (4.06–4.81, mean 4.51) do not correlate reliably with age, sex, or other demographic variables (Section 3.3). Although the youngest two patients and the oldest two cluster at the threshold extremes, the middle of the cohort shows no monotonic pattern, and same-age opposite-sex pairs (CHB-01 and CHB-02) had nearly identical thresholds. This pattern validates the fundamental premise of the patient-specific approach: normal EEG characteristics are individualized properties that cannot be adequately captured by population-based norms, particularly in pediatric populations where baseline patterns change with developmental stage [19], [20], [23].

Traditional seizure detection algorithms using population-based normative data face a conceptual limitation in this context. A universal threshold derived from the cohort mean (4.51) would systematically misclassify epochs in patients with higher (CHB- 08, CHB-10) or lower (CHB-15, CHB-18) individual thresholds, generating both missed seizures and false positives depending on which side of the patient’s true baseline the universal threshold falls. The patient-specific approach eliminates this trade-off by calibrating to each individual’s baseline.

### 4.2 Comparison to existing approaches

At 94.4% event-level sensitivity, the e-norms method exceeds published human expert performance for scalp EEG seizure detection. Inter-rater agreement among board-certified EEG readers typically ranges from 50 to 76% [15], [16], and continuous-monitoring sensitivity can be as low as 20–50%. The method also compares favorably to recent automated systems: Vandecasteele et al. [26] reported 64.1% sensitivity using behind-the-ear EEG in 12 adults with epilepsy, and Alhaskir et al. [27] reported 66.6% overall sensitivity (rising to 100% in a responder subgroup) using wearable ECG in 236 adults with focal-onset impaired-awareness seizures. Munch Nielsen et al. [28] achieved 100% sensitivity in a 12-patient adult pilot study using multimodal recording (EEG plus accelerometer plus ECG), but at the cost of multimodal hardware. Our approach achieves comparable or superior sensitivity using scalp EEG alone, requiring only seizure-free recordings from the target patient — no labeled seizure data, no multi-patient training cohorts, no deep learning. More broadly, related QEEG-clustering approaches are emerging in clinical epilepsy, including EEG dynamics during sedation withdrawal in status epilepticus [29].

A key advantage of the e-norms approach is methodological transparency. The composite stability metric is computed from four interpretable signal features with fixed weights, and threshold derivation follows explicit statistical rules. Unlike deep learning models where detection criteria are opaque, every aspect of the e-norms pipeline can be inspected and clinically interpreted — important for clinical adoption and regulatory approval.

### 4.3 Clinical implications

The 94.4% event-level sensitivity positions this methodology for clinical deployment as a screening and retrospective-review tool in epilepsy monitoring units (EMUs). In typical EMU workflow, continuous EEG generates data volumes that clinicians cannot comprehensively review in real time. The e-norms approach automatically identifies candidate seizure epochs for expert review, enabling clinicians to focus on the highest-yield intervals rather than reviewing hours of unremarkable EEG. In practice, the method would be applied retrospectively at the end of each recording shift or daily review cycle: the patient-specific threshold flags candidate epochs in the preceding interval, and the reviewing neurophysiologist verifies each flagged epoch in the original EEG record. This positions the method as complementary to real-time alarm systems, which prioritize specificity to limit alarm fatigue. The framing of this work is deliberate: it targets sensitivity as a screening problem, not alarm rate as a real-time alarm-device problem, where the cost-benefit balance is fundamentally different.

Several features support clinical readiness. First, the method detected the briefest annotated seizures (6 seconds, CHB-16) at 100% epoch-level sensitivity, performance on events that are challenging for both human readers and automated systems. Second, 100% event-level sensitivity was achieved in 8 of 10 patients, with the two partial-failure cases attributable to characterizable failure modes (Section 4.5) rather than indeterminate detection error. Third, the method handled structural variation across recordings — non-contiguous file numbering and CHB-17’s multi-session structure — without manual intervention.

Implementation requires acquisition of seizure-free baseline EEG, readily obtainable during the early hours of any EMU admission (typical admission: 3–7 days). Baseline sizes in this study ranged from 13 to 36 hours per patient, achievable within one to two days. Once the patient-specific threshold is derived, all subsequent recording from that admission can be screened against it. Computing composite stability for each 2-second epoch and comparing to threshold is computationally trivial for modern systems.

### 4.4 AI-enabled research paradigm

The human-AI collaboration model used in this work (Section 2.4) provided access to cross-disciplinary technical expertise — biostatistics, signal processing, and software engineering — that would otherwise require formal multi-specialist coordination.

This approach enabled a single investigator with deep clinical neurophysiology expertise to systematically apply biostatistical, signal-processing, and software-engineering techniques to a complete cohort analysis.

### 4.5 Limitations, observed failure modes, and future directions

Several limitations warrant consideration. First, the cohort is small (n = 10), pediatric, and from a single institution (CHB-MIT). Generalization to adults, varied epilepsy syndromes, and multi-center data requires further validation; the Siena Scalp EEG Database (PhysioNet) is the next planned step.

Second, this study reports sensitivity but does not characterize false-positive performance, consistent with the screening and retrospective-review framing. Review burden in non-seizure intervals is a separate question for subsequent work.

Third, two patients (CHB-15, CHB-18) showed partial event-level failures distributed across two characterizable failure modes (Section 3.5). The first mode, state-dependent baseline variability, was observed in two of three CHB-15 misses (chb15_06, chb15_10): the patient’s per-file plateau values varied substantially across recordings, leaving the cohort-aggregated threshold too high relative to the local baseline state. A supplementary analysis confirmed that thresholds derived from local windows of surrounding baseline files recovered both events at event level, suggesting that for patients with substantial between-file plateau heterogeneity, windowed thresholding could improve detection without altering the underlying approach.

Fourth, the second mode, observed in chb15_17 and chb18_30, reflects ictal patterns whose composite stability does not exceed the patient’s plateau range — events that cannot be detected by any plateau-derived threshold and represent a limitation intrinsic to single-metric calibration. For chb18_30, seizure-period composite stability was at or below plateau throughout; without raw EEG access from the CHB-MIT EDFs we cannot distinguish among annotation timing offset, atypical ictal pattern, recording-specific artifact, or genuine algorithmic limitation. This ambiguity is intrinsic to algorithm development on annotated EDF data without contemporaneous clinical context, and reinforces the framing of patient-specific composite stability as a screening tool that flags candidate events for visual review rather than a stand-alone diagnostic.

Fifth, the study used retrospective analysis; prospective real-time validation is needed to confirm equivalent performance in streaming clinical settings.

Sixth, the 2-second epoch analysis introduces inherent detection latency. Applications requiring faster response (e.g., closed-loop neurostimulation) would need adaptation to sub-second epochs.

Seventh, the methodology has not been directly benchmarked against other published algorithms on the same CHB-MIT patients, which would strengthen comparative performance claims.

Future validation should address, in priority order: (1) adult cohort validation, (2) prospective real-time validation, (3) head-to-head benchmarking on shared CHB-MIT patients, (4) windowed-threshold investigation for patients with state-dependent baseline variability, (5) review-burden characterization in non-seizure intervals, (6) extension to remaining CHB-MIT patients, and (7) e-norms identification of subclinical epileptiform activity predictive of future seizures.

## 5. Conclusion

This work validates patient-specific e-norms methodology for automated EEG seizure detection across a 10-patient pediatric cohort spanning ages 3 to 18 years. The method achieves 94.4% event-level sensitivity (68 of 72 seizures) and 81.5% epoch-level sensitivity (2,204 of 2,705 epochs) across 263.92 hours of baseline EEG, with 8 of 10 patients reaching 100% event-level sensitivity and the remaining two showing partial failures attributable to characterizable failure modes. These results exceed published human expert inter-rater agreement (50–76%) and recent automated approaches in adult cohorts using behind-the-ear EEG and wearable ECG, while requiring only seizure-free recordings from the target patient and no labeled seizure training data.

The validation demonstrates several clinically important properties: reliable event-level detection across the full annotated seizure-duration ranges from 6 to 264 seconds (a 44-fold range), 100% event-level sensitivity in 8 of 10 patients with two characterizable failure modes accounting for the remaining cases (Section 4.5), and uniform application across the cohort without per-patient parameter adjustment. Patient-specific thresholds (range 4.06–4.81) are essential; no universal threshold could achieve comparable performance given inter-individual variation uncorrelated with demographics.

Beyond seizure detection, this work advances e-norms methodology itself by demonstrating successful application to engineered composite features derived from complex multi-channel neurophysiological signals. This methodological extension — termed *e-norms+* — stands to open e-norms to any domain where meaningful composite features can be constructed from high-dimensional physiological data. The methodology is ready for extended validation in adult populations, prospective clinical deployment, and head-to-head benchmarking against existing automated detection systems.

## Acknowledgements

Ethics and patient consent statement

This study involved secondary analysis of the publicly available, de-identified CHB-MIT Scalp EEG Database (PhysioNet) that was collected under Institutional Review Board approval at Children’s Hospital Boston. As this work involved only retrospective analysis of previously collected, de-identified, publicly available data, no additional ethical approval or patient consent was required.

## Data availability statement

This study used the publicly available CHB-MIT Scalp EEG Database (PhysioNet, https://physionet.org/content/chbmit/1.0.0/). The composite stability metric (Section 2.2) and patient-specific threshold derivation (Section 2.3) are specified in sufficient detail to enable reimplementation by interested investigators.

## Contributions

This work was not performed as part of the author’s academic affiliation with Tufts University School of Medicine. Joe F Jabre is the sole author and was responsible for all aspects of this work including conceptualization, e-norms methodology development, data analysis, validation, interpretation of results, and manuscript preparation. AI collaboration is detailed in the Declaration of generative AI below.

## Declaration of generative AI and AI-assisted technologies in the manuscript preparation process

During the preparation of this work the author used Claude (Anthropic) for computational analysis, statistical calculations, data processing, quality assessment, weighting scheme development, and performance validation. All clinical hypotheses, methodological decisions, result interpretations, and conclusions were the sole responsibility of the investigator who developed the e-norms method, conceptualized the patient-specific approach of the method, specified all methodological requirements, reviewed and validated all analyses, provided clinical interpretation, and made all evidence-based decisions. AI served as analytical tool and collaborator, not as an autonomous decision-maker. After using this tool/service, the author reviewed and edited the content as needed and takes full responsibility for the content of the published article.

